# Converting Passive Filtration Media into Active Air Biofiltration Surfaces for Airborne Viral Reduction

**DOI:** 10.64898/2026.04.29.26352113

**Authors:** Ralph G. Dacey, Chukwudi C. Ezeala, Tom Kennedy, Emily Agnello, Alexander Ribbe, Sérgio Cavaleiro, Nelson Marques, Carly Emamjoneh, Robert Roth

## Abstract

Conventional air filtration relies on passive mechanical capture without pathogen inactivation, where viral reduction must be balanced with airflow and energy performance. We developed an Ablative Polymer Coated (APC) filtration system that converts passive filters into active pathogen-reducing surfaces while maintaining low airflow resistance. Unlike conventional approaches requiring denser, higher-resistance media, this strategy enhances biological performance at the filter surface without equivalent aerodynamic penalties. The coating incorporates benzalkonium chloride within a polyvinyl acetate/acrylate matrix for controlled ablative exposure. Performance was evaluated using transmission electron microscopy (TEM), aerosol challenge testing, and HVAC-scale filtration. Ablative exposure caused progressive structural disruption of MS2 bacteriophage, the SARS-CoV-2 simulant. In aerosol challenge testing, coated media achieved up to 99.997% viral filtration efficiency under respiratory airflow conditions. In HVAC (Heating, Ventilation, and Air Conditioning)-scale testing, coated filters achieved >85% viral filtration efficiency with minimal pressure-drop increase. Computational fluid dynamics modeling confirmed uniform airflow distribution without significant turbulence generation. Energy analysis suggested coated filters may reduce energy demand relative to conventional higher-resistance configurations while improving biological performance. These findings support ablative polymer-coated media as a strategy for reducing airborne viral burden without aerodynamic penalties of higher-efficiency passive filtration, suggesting an approach that complements rather than depends solely on tighter filter design.

## Introduction

Respiratory pathogens present ongoing challenges across indoor environments, with SARS-CoV-2, influenza viruses, and agricultural pathogens such as PRRSV (Porcine reproductive and respiratory syndrome virus) and HPAI (Highly Pathogenic Avian Influenza), maintaining viability in aerosol form while circulating through ventilation systems for extended periods [1–3]. The COVID-19 pandemic, seasonal influenza, and PRRSV outbreaks have demonstrated the substantial economic impact of airborne pathogens, highlighting the need for control strategies that achieve both high biological effectiveness and practical implementation feasibility.

Current filtration approaches face fundamental limitations restricting widespread adoption. Standard HVAC filters capture particles mechanically but provide no pathogen inactivation, with viruses potentially remaining viable on filter fibers for hours or days [4,5]. Higher biological filtration efficiency has traditionally been achieved through denser filter media, creating substantial pressure drops and increasing fan energy consumption by 200–300% when upgrading from MERV (Minimum Efficiency Reporting Value) 8 to MERV 15 filters [6,7]. Agricultural facilities requiring high ventilation rates face challenges, as high-efficiency filtration often exceeds existing fan capacity despite pathogen outbreaks causing substantial production losses.

Current air filtration standards compound these limitations, particularly for submicron particles relevant to viral transmission. The widely used ANSI/ASHRAE (American Society of Heating, Refrigerating and Air-Conditioning Engineers) 52.2 standard evaluates filter efficiency only for particles ranging from 0.3 to 10.0 μm, excluding the nanoparticle size range where many respiratory viruses operate [8]. Recent research indicates that “current knowledge of filter performance for nanoparticles is still very limited,” highlighting fundamental gaps in filtration effectiveness against biological pathogens [8]. The importance of addressing airborne transmission has been emphasized by foundational work on airborne contagion [9] and recent studies confirming the airborne nature of SARS-CoV-2 transmission [10–12].

Existing air filtration technologies fall into three main categories. However, each approach has significant limitations: passive mechanical filters requiring increasingly dense media for higher efficiency, creating substantial energy penalties [13–15]; active systems like UV-C or plasma that add complexity and maintenance burden [16,17]; and ablative treatments that either compromise airflow or lack proven viral efficacy under realistic conditions [18–20]. These limitations demonstrate fundamental tradeoffs in achieving high viral filtration efficiency (>85% VFE), which typically require substantial energy penalties (>0.45 in w.g. pressure drop) and complex installation requirements.

These constraints have increased interest in alternative approaches that improve biological performance without depending solely on mechanical filtration density or complex active systems. Ablative polymer-coated filtration media represents a promising strategy that incorporates ablative compounds into a polymer matrix applied to fibrous filtration media, potentially combining particle capture with pathogen-reducing surface activity [21–24]. Unlike conventional approaches targeting improved capture through denser media or add-on active systems, this strategy aims to enhance biological performance at the filter surface without imposing equivalent aerodynamic and energy penalties.

In this study, we evaluated ablative polymer-coated filtration media under conditions relevant to aerosol exposure and ventilation operation. Using computational fluid dynamics modeling (CFD) [25–27], transmission electron microscopy, aerosol challenge testing, and HVAC-scale filtration measurements, we examined whether the coating application alters modeled particle transport behavior, disrupts viral structural integrity, reduces downstream recovery of infectious virus, and accomplishes these effects without introducing detectable emissions or imposing substantial airflow resistance. We hypothesized that ablative polymer-coated media would provide improved viral reduction relative to uncoated controls while maintaining low airflow resistance, supporting a filtration strategy distinct from conventional higher-resistance passive filter upgrades or complex active treatment systems.

## Materials and Methods

### Study design overview

This study evaluated Ablative Polymer-Coated (APC) filtration media as a platform for active air biofiltration under conditions relevant to aerosol exposure and ventilation operation. The experimental design integrate CFD modeling, transmission electron microscopy, aerosol challenge testing, ventilation-relevant single-pass filtration testing, pressure drop characterization, energy performance analysis, and environmental emissions testing to assess whether coating application altered modeled particle transport behavior, disrupted viral structural integrity, reduced downstream recovery of infectious virus, improved higher-flow filtration performance, and accomplished these effects without introducing detectable emissions of selected target analytes or imposing substantial airflow resistance.

## Materials

### Coating Materials and Preparation

The ablative polymer coating consists of a copolymer matrix with EHEC rheology modifier/binder and benzalkonium chloride as the active antimicrobial agent. While specific formulation ratios and processing parameters constitute proprietary information currently under patent protection, the complete coating formulation and application protocol will be made available to qualified researchers for academic research purposes through a material transfer agreement. Requests should be directed to the corresponding author (Ralph G. Dacey Jr.) or directly to BioActive Technology LLC. Commercial applications may require separate licensing arrangements.

#### Filter Substrates

Commercial fibrous filtration media included 5W100 (MERV 8), SP-100 (MERV 10), XP-100, 1" PE, and 2" PE substrates obtained from commercial suppliers. Additional MERV 7 and MERV 10 class substrates were used for validation of experiments.

#### Ablative Coating Components

The three-component polymer formulation consisted of: (1) polyvinyl acetate/2-ethylhexyl acrylate copolymer (polymeric matrix), (2) ethyl hydroxyethyl cellulose (EHEC, rheology modifier and binder), and (3) benzalkonium chloride (BAC, active antimicrobial agent). All chemical components were obtained from commercial suppliers and used as received.

#### Viral Challenge Organism

Bacteriophage MS2 (ATCC 15597-B1) was selected as the viral surrogate because it is a well-established, conservative model organism for aerosol filtration, transmission, and antimicrobial surface research, and has been used extensively as a simulant for SARS-CoV-2 [28,29]. As a small (∼27 nm), non-enveloped, icosahedral positive-sense single-stranded RNA virus, MS2 possesses high environmental stability and is notably more resistant to quaternary ammonium compounds such as benzalkonium chloride than enveloped respiratory viruses including SARS-CoV-2, influenza, and PRRSV [30,31]. This resistance profile makes MS2 a conservative surrogate for evaluating antiviral efficacy, as successful inactivation of MS2 suggests even greater efficacy against more susceptible enveloped viruses [32].

### Ablative Polymer Coating Preparation

Ablative coatings were prepared using a three-component polymer formulation consisting of: (1) a polyvinyl acetate/2-ethylhexyl acrylate copolymer providing the polymeric matrix for substrate adhesion and mechanical properties, (2) ethyl hydroxyethyl cellulose (EHEC) serving as a rheology modifier and coating binder, and (3) benzalkonium chloride (BAC) as the active ablative component (Fig 1). The cellulose-derived additives were incorporated to modulate coating behavior and regulate exposure of the active agent at the media surface. The polymer formulation was applied to fibrous filtration media using two distinct methods depending on the test series: spray-coating and dip-coating approaches. For spray-coating applications, the polymer solution was applied using controlled spray parameters to ensure uniform distribution across the filter substrate. For dip-coating applications, filter substrates were immersed in the coating solution under controlled conditions to achieve consistent coating thickness. Coated materials were dried under controlled temperature and humidity conditions before structural characterization, aerosol testing, and airflow performance evaluation.

**Figure 1:**
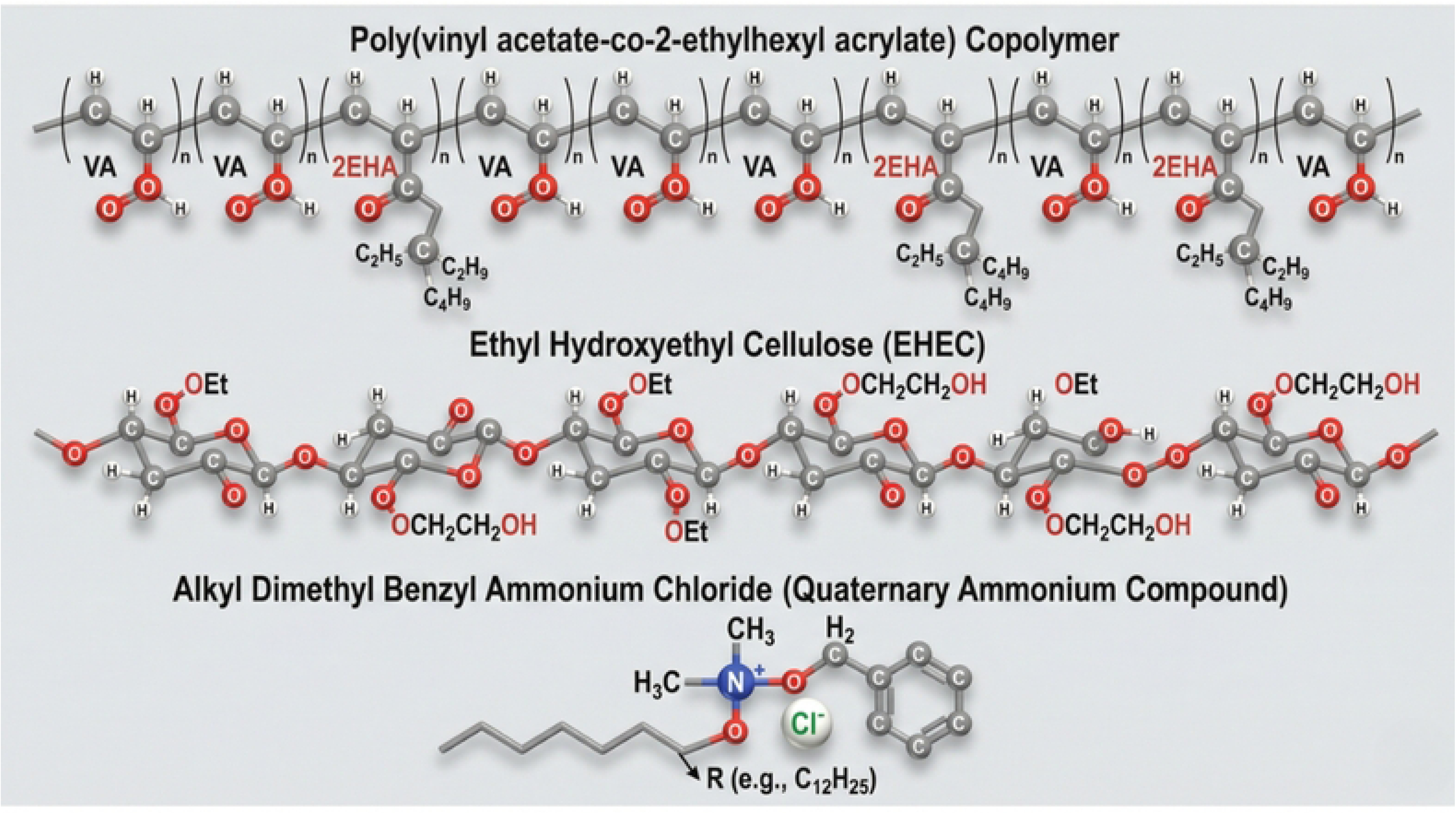
Chemical composition of the three-component ablative polymer coating system. (Top) Poly(vinyl acetate-co-2-ethylhexyl acrylate) copolymer backbone showing alternating vinyl acetate (VA) and 2-ethylhexyl acrylate (2EHA) monomeric units. This copolymer provides the primary polymeric matrix for substrate adhesion and mechanical properties. (Middle) Ethyl hydroxyethyl cellulose (EHEC) structure displaying the modified cellulose backbone with ethyl (OEt) and hydroxyethyl (OCH_2_CH_2_OH) substituents. EHEC functions as a rheology modifier and coating binder. (Bottom) Alkyl dimethyl benzyl ammonium chloride, a quaternary ammonium compound (QAC) with a variable alkyl chain length (R, typically C_12_H_25_), serving as the active ablative agent. The multi-component formulation enables both strong fiber adhesion through the polymer matrix and sustained ablative activity through the surface-active QAC.

### Transmission Electron Microscopy

Transmission electron microscopy (TEM) experiments were performed to evaluate structural changes in viral morphology following ablative exposure. Bacteriophage MS2 particles (ATCC, 15597-51) were used as a viral surrogate model, as is common for aerosol filtration and antiviral surface studies. MS2 particles suspended in 1x PBS were either added to a 1 % polymer solution, or added to polymer-enhanced filter fragments, followed by 10 min sonication and aspiration of the resulting solution. Control samples included untreated viral particles and particles exposed to 1% benzalkonium chloride in the same manner as above. 3.5 uL of sample were applied to a 400-mesh copper grid with carbon film and stained using a single-drop method with 1% Uranyl Acetate. Samples were imaged using a ThermoFisher Tecnai T12 transmission electron microscope operating at 120kV with a LaB6 filament. Images were acquired at a nominal magnification of 52,000x on a TVIPS 2k CMOS camera with a 500ms exposure time, and analyzed at the University of Massachusetts Institute for Applied Life Sciences.

### Aerosol Challenge Testing

Aerosol challenge experiments were conducted at Nelson Laboratories using MS2 bacteriophage as the challenge organism under respiratory-relevant conditions. The experimental design evaluated four coating concentrations (1.25%, 2.5%, 5%, and 10% w/w) plus untreated controls, with three replicate experiments performed per coating concentration (n=3 per group).

Aerosolized MS2 particles were generated at approximately 3.0 ± 0.3 μm mean diameter and passed through test media at 20 L min⁻¹ flow rate for 1 minute under controlled environmental conditions. Challenge aerosols were generated using standardized protocols to ensure consistent particle size distribution and viral concentration across experimental conditions. Infectious virus recovered downstream of the filtration media was quantified by plaque-forming unit (PFU) assay using standard microbiological techniques.

Viral filtration efficiency (VFE) was calculated using the formula: VFE = (1 - [PFU_downstream/PFU_challenge]) × 100%, where PFU_challenge represents the viral load in the challenge aerosol and PFU_downstream represents the recoverable infectious virus collected downstream of the test medium.

Statistical analysis of aerosol challenge data employed one-way analysis of variance (ANOVA) to compare viral filtration efficiency across coating concentrations. Tukey’s Honestly Significant Difference (HSD) test was applied for pairwise comparisons between concentration groups when ANOVA indicated significant differences (p < 0.05). Experimental assumptions were verified through normality assessment using the Shapiro-Wilk test and homogeneity of variance evaluation using Levene’s test. Effect size was calculated using eta-squared (η^2^) to determine the proportion of variance in VFE explained by coating concentration.

### Ventilation-Scale Filtration Testing

Higher-flow single-pass filtration testing was performed by LMS Technologies using aerosolized MS2 bacteriophage challenge under conditions representative of HVAC ventilation operation.

Testing was conducted at 819 CFM under ASHRAE 52.2-compliant conditions using fibrous filtration media representative of mid-MERV applications. Test substrates included 5W100 (MERV 8) and SP-100 (MERV 10) commercial filter media, with additional validation performed on MERV 7 and MERV 10 class substrates. Each substrate was evaluated in both untreated control and ablative-coated conditions, with coatings applied using both spray and dip-coating methods to assess application technique effects. Three replicate experiments were conducted per condition (n=3) to ensure statistical reliability.

In the LMS single-pass testing system, viral removal efficiency and simultaneous pressure drop measurements were obtained under airflow conditions representative of commercial ventilation operation. Additional testing was performed at 295 fpm face velocity for selected media combinations to evaluate performance under varying operational conditions.

Energy-dispersive X-ray spectroscopy (EDS) was employed in related optimization experiments to assess coating localization and distribution on fiberglass fiber surfaces, confirming that the ablative polymer coating was successfully deposited along individual filter fibers without disrupting the overall porous filter architecture.

Statistical analysis of single-pass filtration data employed two-sample t-tests for comparing treated versus control conditions, with Cohen’s d calculations for effect size assessment. Multiple comparisons were applied across different substrate types to evaluate coating effectiveness across various commercial filter media.

### Pressure Drop Characterization

Independent pressure drop analysis was conducted across multiple filter substrates to evaluate aerodynamic penalties associated with ablative coating application. Testing was performed under standard conditions (70°F temperature, 40% relative humidity) using ASHRAE-compliant procedures to ensure reproducible and industry-relevant measurements.

Measurements were taken across a range of flow rates from 205 to 1024 CFM, with primary analysis conducted at 819 CFM to provide direct comparison with viral filtration test conditions. Test substrates included SP-100, XP-100, 1" PE, and 2" PE filter media to assess coating effects across different substrate materials and configurations.

Percentage changes in pressure drop were calculated using the formula: % Change = [(P_coated - P_control)/P_control] × 100%, where P_coated represents the pressure drop across coated media and P_control represents the pressure drop across equivalent uncoated control media.

#### Important Technical Note

All pressure drop measurements were conducted on flat media samples to ensure standardized comparison conditions across different substrates and coating applications. Commercial HVAC filters are typically pleated rather than flat configuration, and industry experience indicates that pleating typically reduces pressure drop by approximately 50% compared to equivalent flat media due to increased effective surface area and improved airflow distribution patterns. Therefore, the pressure drop values reported in this study represent conservative estimates for practical implementation. Actual field performance of pleated coated filters would likely demonstrate even lower pressure penalties than those measured under standardized flat-media test conditions.

### Energy Performance Analysis

Energy performance interpretation was based on the quantitative relationship between filtration efficiency improvements, pressure drop characteristics, and ventilation system practicality rather than comprehensive whole-building energy modeling. The analysis framework evaluated whether improved viral removal performance could be achieved without the substantial fan-energy penalties typically associated with conventional dense passive filtration upgrades.

Because the ablative-coated media were specifically designed to improve biological performance without materially increasing airflow resistance, the energy-relevant evaluation focused on whether the observed filtration efficiency gains could be accomplished while maintaining pressure drop characteristics similar to baseline uncoated filter substrates. This interpretation was supported qualitatively by the observed pressure-drop stability in higher-flow single-pass filtration tests and validated through independent pressure drop characterization across multiple substrate types and operational flow rates.

### Computational Fluid Dynamics

Computational fluid dynamics (CFD) simulations were performed by FS Dynamics to compare modeled particle transport behavior in coated and uncoated porous filter media under controlled numerical conditions. Three-dimensional filter geometries were reconstructed from shared computed tomography (CT) scan DICOM files and processed using 3D Slicer version 5.6.2 software for geometry preparation and Star-CCM+ v19.02.013 (Siemens PLM Software) for computational analysis.

The computational mesh consisted of 12,725,740 control volumes for the coated medium analysis, with corresponding mesh density applied to uncoated control geometries to ensure numerical consistency between comparative simulations. The CFD model was used to evaluate velocity field distributions, individual particle trajectory behavior, flow tortuosity characteristics, traversal time analysis, and pressure drop predictions in both coated and uncoated media configurations under matched boundary conditions and flow parameters.

Statistical comparisons between coated and uncoated media included detailed tortuosity analysis calculated as τ = L_path/L_strait, where L_path represents the actual particle pathline length through the porous medium and L_strait represents the straight-line inlet-to-outlet distance.

Traversal time analysis was nondimensionalized as τ_trav = (d/u_in) × t, where d represents filter depth, u_in represents inlet velocity, and t represents transit time through the porous medium.

Pressure drop measurements were reported in mPsi with comparative analysis between reconstructed coated and uncoated geometries. Particle retention statistics were calculated from particle trajectory simulations, with retention percentages based on the ratio of particles captured within the porous medium to particles released at the domain inlet.

Median values were calculated for both tortuosity and traversal time distributions due to non-normal distribution characteristics evident in cumulative distribution function analysis of the particle trajectory datasets. Because the CFD analysis relied on reconstructed geometries derived from imaging data rather than direct in-situ flow measurements, results were interpreted as supportive mechanistic modeling rather than definitive quantitative predictions of in-filter antiviral mechanisms.

### Environmental Emissions Testing

Comprehensive environmental emissions testing was conducted by TRC Environmental Corporation at the Industrial Polymers and Chemicals test facility in Shrewsbury, Massachusetts, during August 29-30, 2023. The testing protocol evaluated potential emissions of benzalkonium chloride and selected volatile organic compounds during controlled filter operation.

Untreated control filters were evaluated on day 1 and ablative-treated filters on day 2 under identical environmental and operational conditions. Testing was performed at 400 CFM airflow rate with simultaneous inlet and outlet monitoring to assess any emissions attributable to the treated filter media.

For volatile organic compound analysis, integrated Summa canister samples were collected at both inlet and outlet locations and analyzed according to U.S. EPA Method TO-15 using gas chromatography-mass spectrometry (GC/MS). Target analytes specifically included vinyl acetate, benzene, and benzyl chloride, with results also reported for the complete TO-15 analyte library to provide comprehensive volatile organic compound screening.

For benzalkonium chloride analysis, air samples were drawn through 37 mm cassette filters and analyzed using a proprietary ALS Laboratories GC/MS method specifically validated for quaternary ammonium compound detection in air matrices. Two 120-minute TO-15 samples and one 8-hour BAC sample were collected at both inlet and outlet locations for each control and treated filter condition to ensure adequate sample collection and analytical reliability.

Environmental conditions during testing were controlled and monitored, with temperature and humidity maintained within specified ranges to ensure consistent test conditions between control and treated filter evaluations. Background contamination was assessed through analysis of inlet air samples to distinguish filter-attributable emissions from ambient environmental contributions.

### Statistical Analysis

All statistical analyses were performed using R version 4.5.3 GUI 1.82 Big Sur Intel build (8587) and RStudio Version 2026.01.2+418 statistical software. Experimental designs employed n=3 replicate experiments per condition with statistical significance threshold set at α = 0.05 for all hypothesis testing.

Aerosol challenge concentration-dependent data were analyzed using one-way analysis of variance (ANOVA) with Tukey’s Honestly Significant Difference (HSD) post-hoc testing for pairwise comparisons between coating concentration groups. Single-pass filtration data comparing treated versus control conditions employed two-sample t-tests with Cohen’s d effect size calculations using pooled standard deviation estimates.

Statistical assumptions were rigorously verified prior to analysis implementation. Normality was assessed using the Shapiro-Wilk test for all datasets, and homogeneity of variance was evaluated using Levene’s test for multi-group comparisons. When assumption violations were detected, appropriate non-parametric alternatives or data transformations were applied as indicated by diagnostic testing results.

Effect sizes were calculated using eta-squared (η²) for ANOVA analyses to determine the proportion of total variance in viral filtration efficiency explained by coating concentration variables. For t-test comparisons, Cohen’s d effect sizes were calculated to quantify the magnitude of treatment effects independent of sample size considerations.

All data is presented as mean ± standard deviation with 95% confidence intervals provided where appropriate. Statistical significance was consistently evaluated at the α = 0.05 level, with exact p-values reported for all hypothesis tests to enable comprehensive interpretation of statistical evidence strength.

## Results

### Existing filtration technologies demonstrate fundamental performance tradeoffs

Current air filtration approaches face a fundamental limitation: high viral efficiency comes at the cost of energy penalties or installation complexity (Table 1). These constraints motivated development of ablative polymer coating technology to achieve biological performance improvements without proportional infrastructure demands.

**Table 1:** Performance characteristics of current air filtration technologies for achieving high viral filtration efficiency. Quantitative comparison demonstrates energy and implementation tradeoffs inherent in existing approaches.

| Technology | VFE (%) | Pressure Drop | Energy Impact | Installation |
| --- | --- | --- | --- | --- |
| MERV 13 (passive) | 75-85<sup>1</sup> | >0.45 in w.g.<sup>2</sup> | High<sup>3</sup> | Retrofit required |
| MERV 15 (passive) | 85-95<sup>1</sup> | >0.9 in w.g.<sup>2</sup> | Very High<sup>3</sup> | Major upgrade |
| HEPA | >99.97<sup>4</sup> | >1.2 in<br>w.g.<sup>5</sup> | Extreme<sup>3</sup> | Full system<br>replacement |
| UV-C Systems | Variable<sup>6</sup> | N/A | Moderate<sup>7</sup> | Complex<br>installation |
| **Ablative Polymer Coating**<br>**(APC) (MERV 10)** | **86.7** | **0.358 in**<br>**w.g.** | **Low** | **Drop-in**<br>**replacement** |

### Computational modeling suggested altered particle transport through coated porous media

The proposed mechanism for enhanced viral reduction involves increased particle-surface interaction opportunities within coated porous media (Fig 2). CFD simulations indicated generally similar transport behavior in coated and uncoated porous media, with a modest shift toward longer traversal times in coated conditions. Approximately 25% of particles in coated medium vs. 10% in uncoated medium exceeded the unimpeded-flow reference condition. The coated medium showed differences in porosity and surface area that may increase particle-surface interaction opportunities. These findings should be interpreted cautiously due to potential geometry-reconstruction effects noted in the CFD analysis.

**Figure 2:**
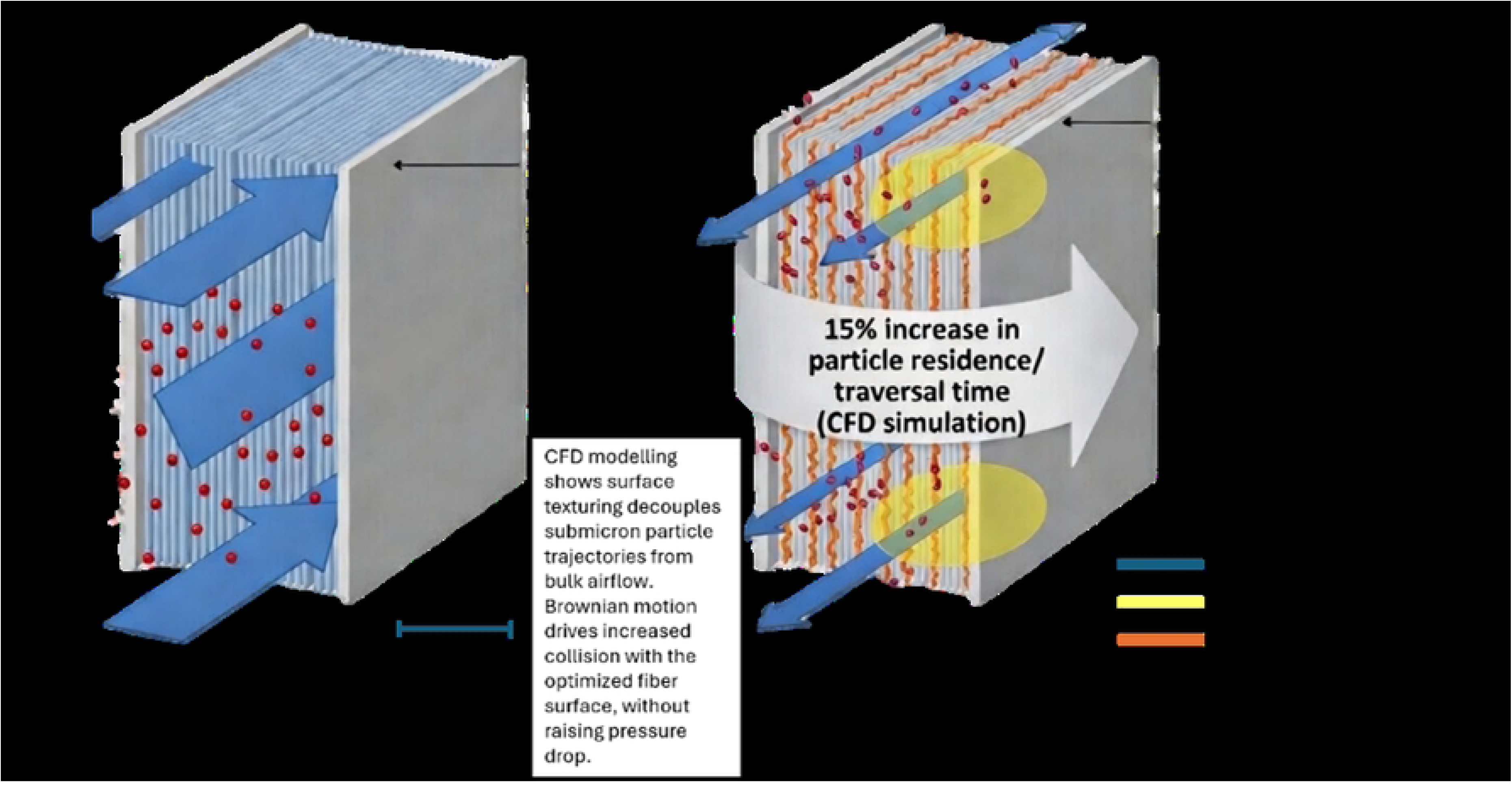
Conceptual schematic of proposed coating-associated effects on particle transport through filtration media. The schematic illustrates a proposed interpretation in which coated media may increase opportunities for particle–surface interaction relative to uncoated media. This figure is conceptual and is intended to aid interpretation of the CFD and filtration results; it does not represent a direct measurement of mechanism

### Transmission electron microscopy revealed progressive viral structural disruption following ablative exposure

Transmission electron microscopy (TEM) was used to examine morphological changes in MS2 bacteriophage following exposure to ablative treatments. MS2 was selected as a surrogate viral model because of its structural stability and widespread use in aerosol, filtration, and antiviral surface studies.

Untreated control particles displayed intact, well-defined capsid morphology consistent with native MS2 structure (Fig. 3b). In contrast, particles exposed to 1% benzalkonium chloride (BAC) showed partial structural damage, including capsid deformation and loss of uniform particle morphology (Fig. 3c). This limited virucidal activity is in line with previous observations that BAC shows a limited inactivation against MS2.^37^More extensive disruption was observed after exposure to the polymer coated filter media, with TEM images revealing pronounced particle collapse, severe capsid damage, and marked loss of structural integrity (Fig. 3d). This observation was conserved whether the viral particles were exposed to polymer coated filter media or captured on the treated filter material (Fig. 3e).

**Figure 3:**
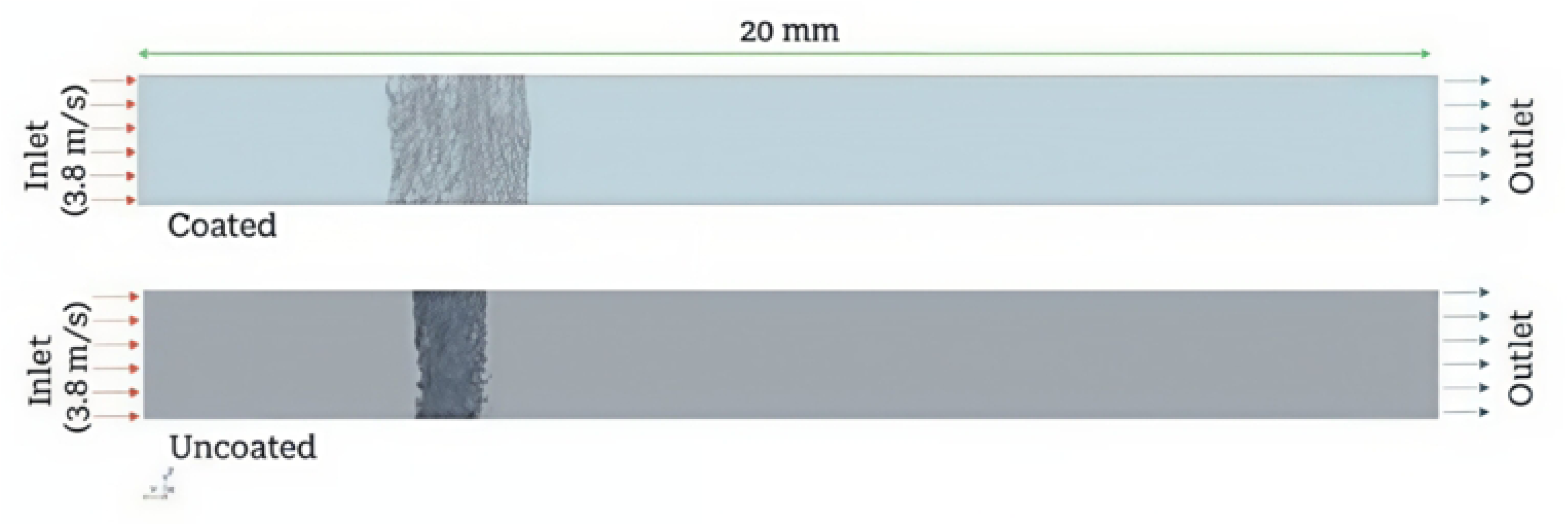
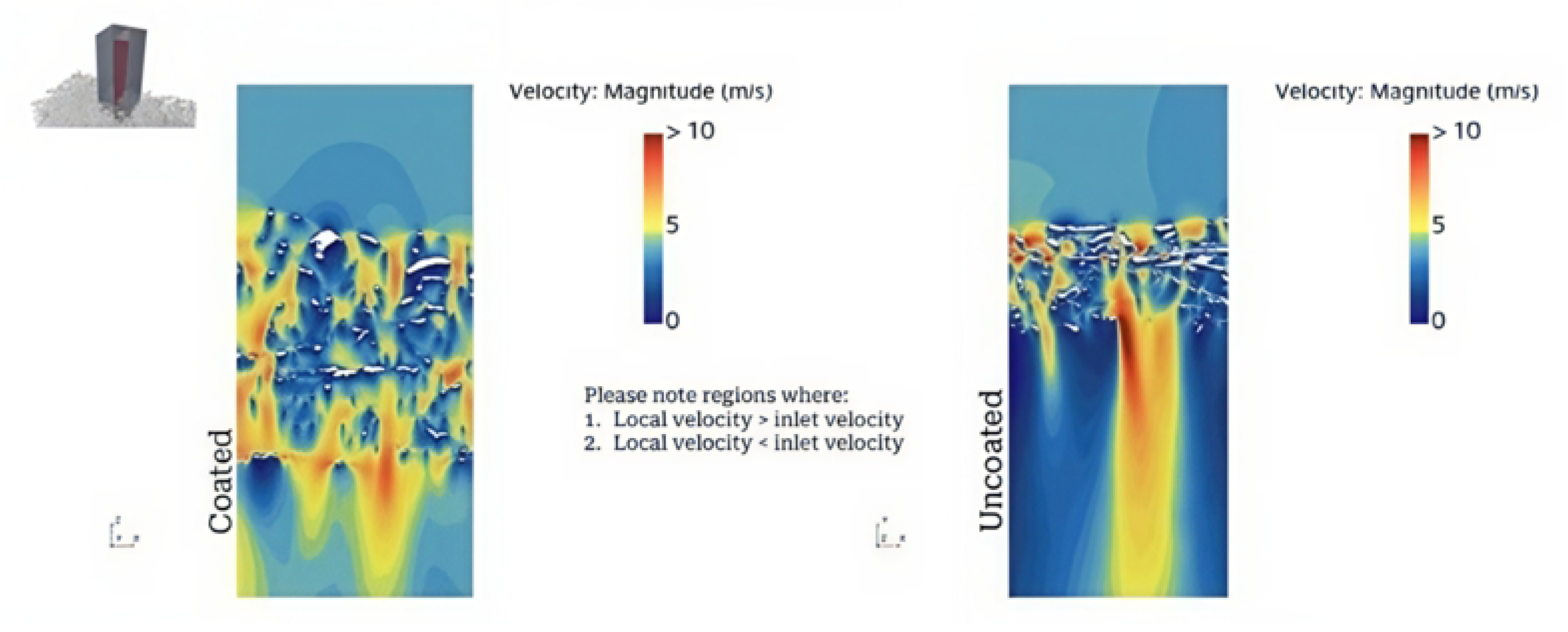
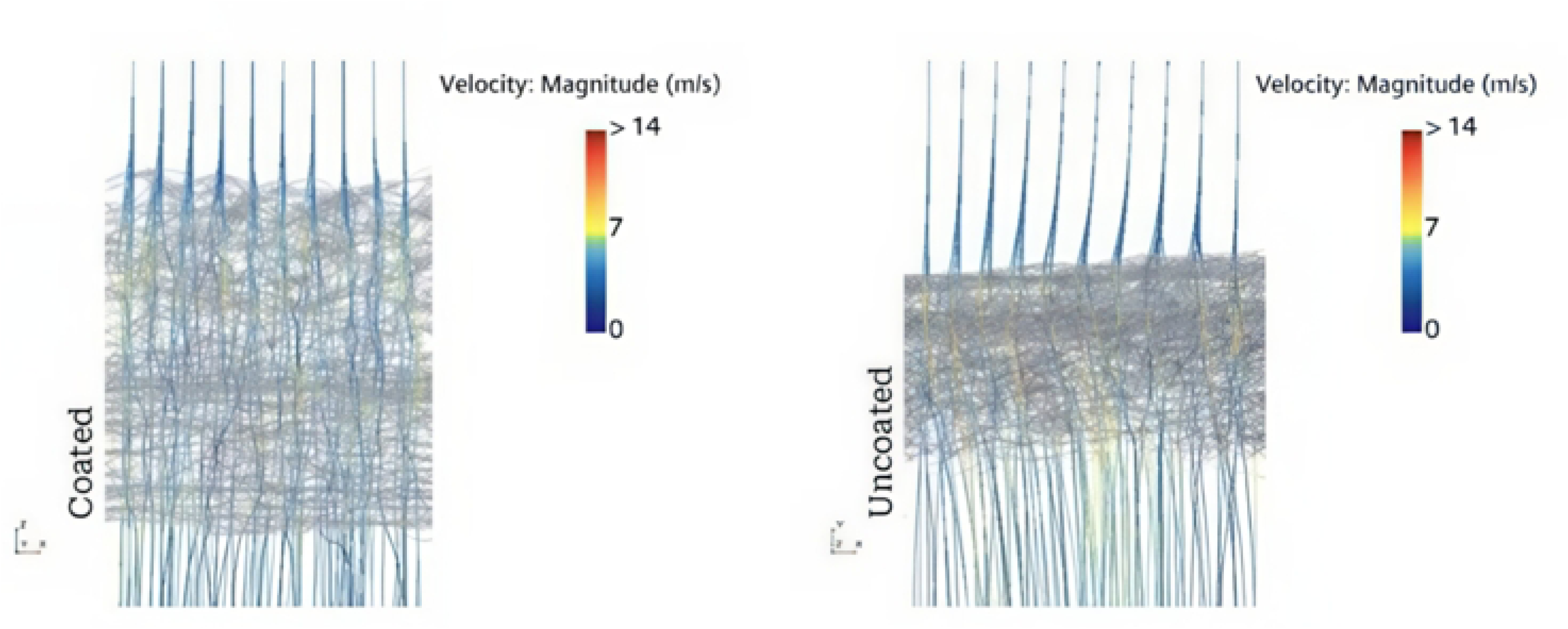
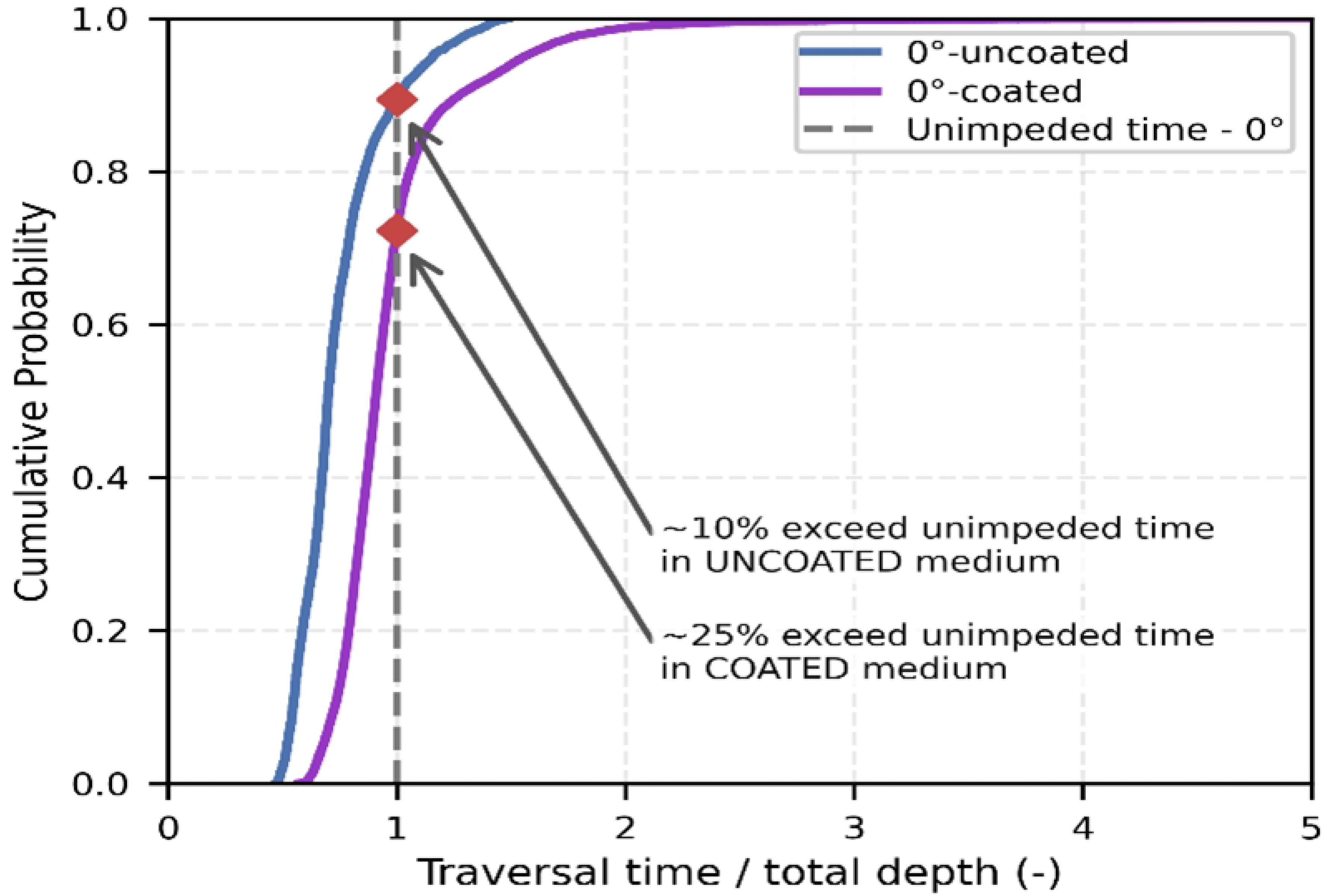
Computational modeling suggested altered particle transport through coated porous media. **a,** CFD model setup showing coated and uncoated porous media within the simulated flow domain. **b,** XZ-plane velocity magnitude comparison for coated and uncoated media. **c,** Representative particle trajectories through coated and uncoated porous media. **d,** Cumulative distribution of filter traversal time showing that a greater fraction of particles in the coated medium exhibited traversal times longer than the unimpeded-flow reference.

**Figure 4:**
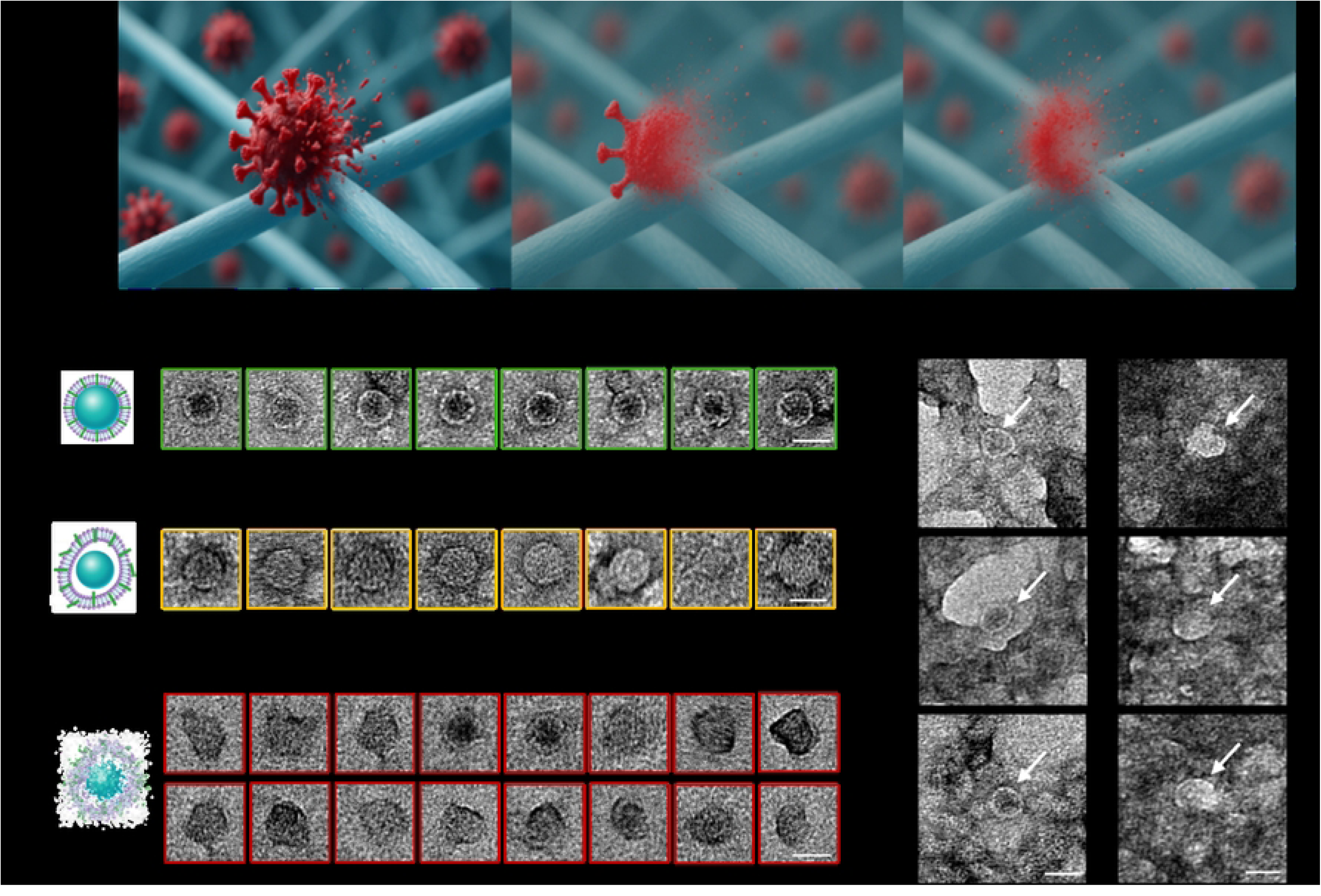
Transmission electron microscopy reveals progressive structural disruption of MS2 bacteriophage following ablative treatment. **a,** Conceptual overview of the proposed inactivation mechanism. Contact with the ablative coating is proposed to promote structural destabilization of the viral capsids, rendering them non-infectious. **b**, Untreated MS2 bacteriophage particles showing intact capsid morphology. **c,** MS2 particles exposed to 1% benzalkonium chloride (BAC), showing partial capsid deformation and structural damage. **d,** MS2 particles exposed to the filter with enhanced ablative polymer coating, showing extensive particle collapse and loss of structural integrity. **e,** Representative micrographs demonstrate a similar level of viral disruption (arrows) when imaged on treated filter (right), as compared to untreated filter (left). TEM imaging was performed at the University of Massachusetts Amherst, Institute for Applied Life Sciences. Scale bars represent 25 nm.

These observations indicate that ablative polymer treatment progressively disrupts MS2 particle structure, with the polymer-coated formulation producing greater damage than BAC alone. A conceptual overview of the proposed inactivation mechanism is shown in Fig. 3a, in which contact with the ablative coating is proposed to promote structural destabilization of viral particles and reduced infectivity.

### Aerosol challenge testing demonstrated a concentration-dependent reduction in infectious viral recovery

Under respiratory-relevant aerosol challenge conditions, untreated control filters recovered approximately 7.7 × 10^5 PFU downstream, corresponding to a baseline VFE of 67.2%. Coating applications produced a strong concentration-dependent reduction in recoverable infectious virus. At 1.25% and 2.5% coating concentrations, downstream recovery decreased to approximately 1.4 × 10^4 PFU and 1.1 × 10^4 PFU, corresponding to VFE values of 94.8% and 95.2%, respectively. At 5% and 10% coating concentrations, downstream recovery fell further to approximately 2.9 × 10^3 PFU and 5.4 × 10^2 PFU, corresponding to VFE values of 99.7% and 99.997%. These results show that increasing coating concentration was associated with progressively lower downstream recovery of infectious MS2.

### Ventilation-relevant testing showed improved viral removal under higher-flow conditions

Single-pass testing under ventilation-relevant conditions demonstrated statistically significant improvements in viral filtration efficiency with minimal aerodynamic penalties (Tables 2-3, Figure 6a). SP-100 coated media achieved 86.70 ± 1.31% VFE compared to 30.80 ± 0.79% for controls, representing a 181% improvement (p < 0.001, Cohen’s d = 51.68) that exceeds MERV 15 standards. The 5W100 substrate similarly showed significant improvement (52.87 ± 1.86% vs. 20.67 ± 1.36% control, p < 0.001, Cohen’s d = 19.77). Dipped application consistently outperformed sprayed application across both substrates, with SP-100 dipped media achieving optimal performance (Figure 6a).

**Figure 5:**
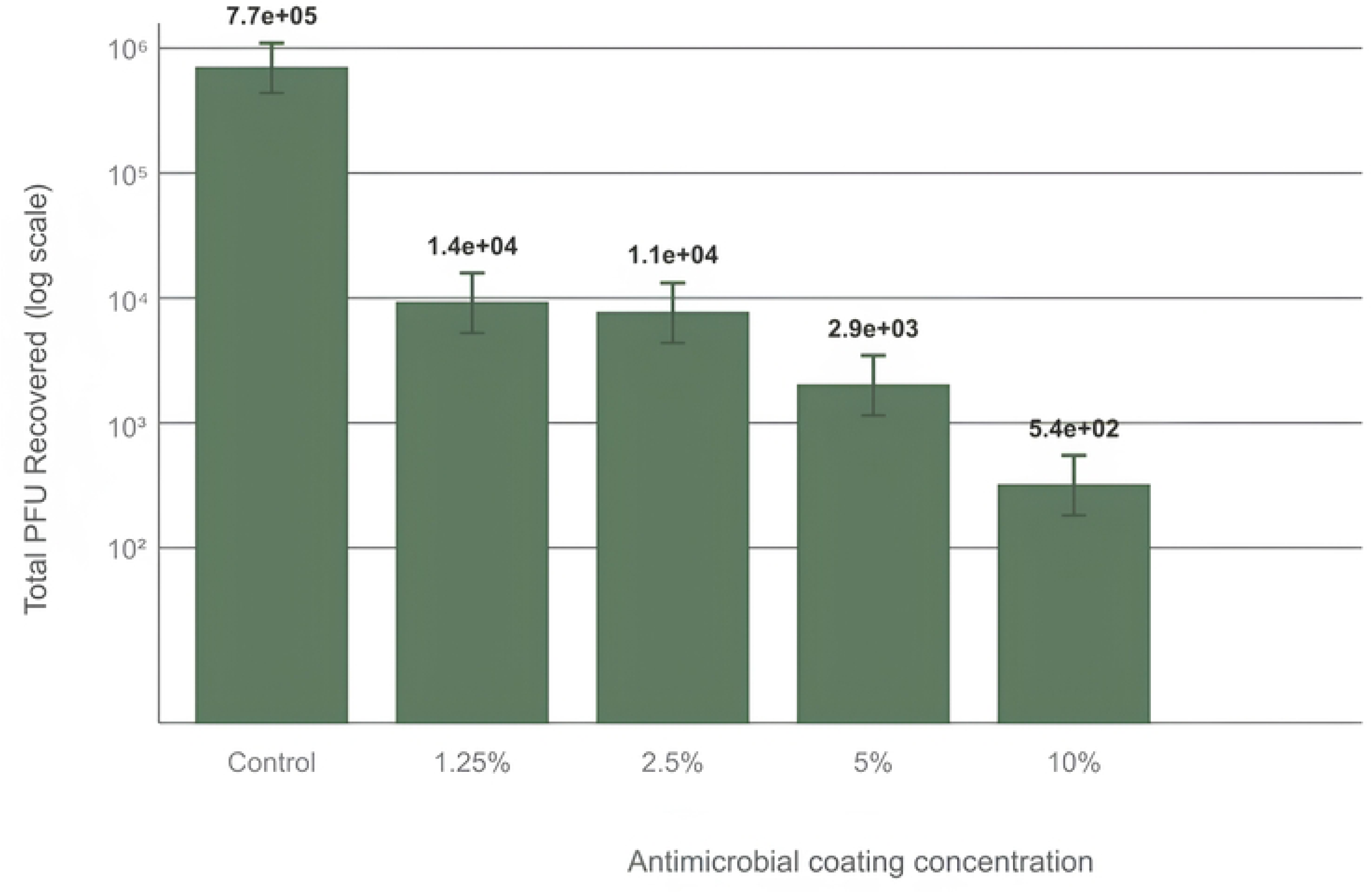
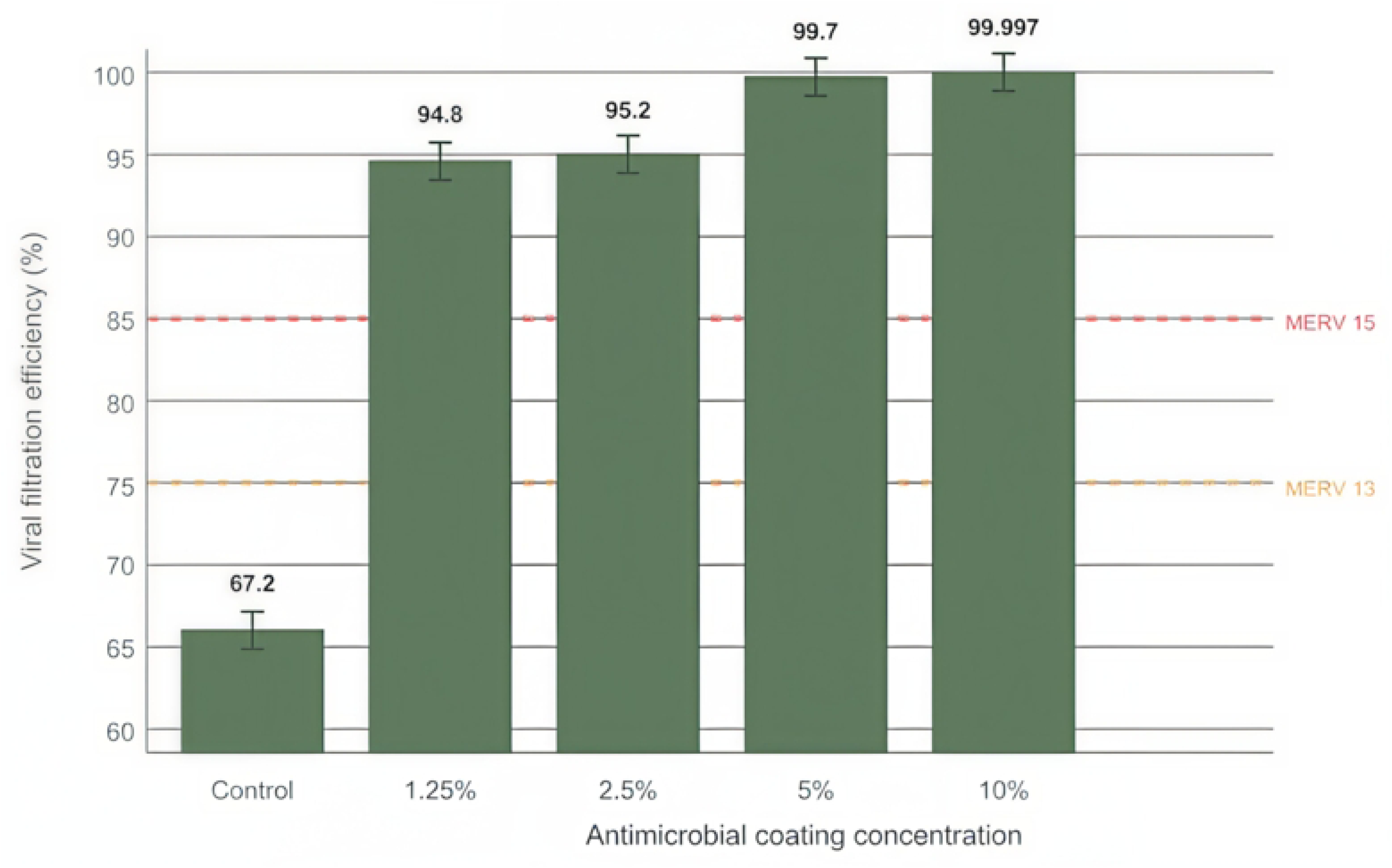
Aerosol challenge testing demonstrated a concentration-dependent reduction in infectious MS2 recovery with increasing antimicrobial coating concentration. **a,** Total plaque-forming units (PFU) recovered downstream of filtration media following aerosol challenge with MS2 bacteriophage. Downstream recovery decreased progressively with increasing ablative coating concentration. **b,** Viral filtration efficiency (VFE) of filtration media as a function of ablative coating concentration. Untreated media showed 67.2% VFE, whereas coated media reached 94.8%, 95.2%, 99.7%, and 99.997% VFE as coating concentration increased. Aerosolized MS2 bacteriophage particles were passed through filtration media under controlled respiratory-relevant airflow conditions, and downstream infectious virus was quantified by PFU assay.

**Figure 6:**
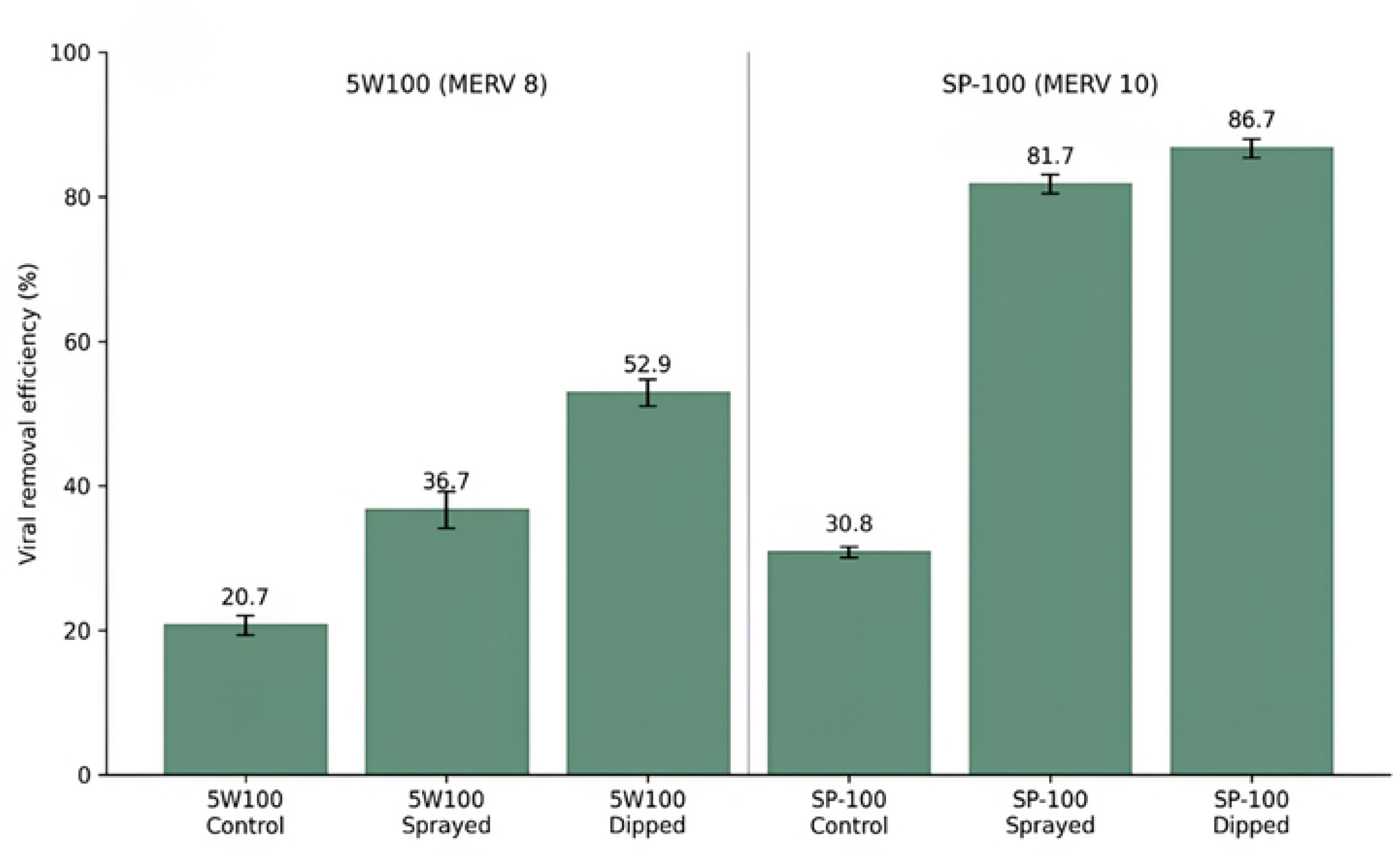
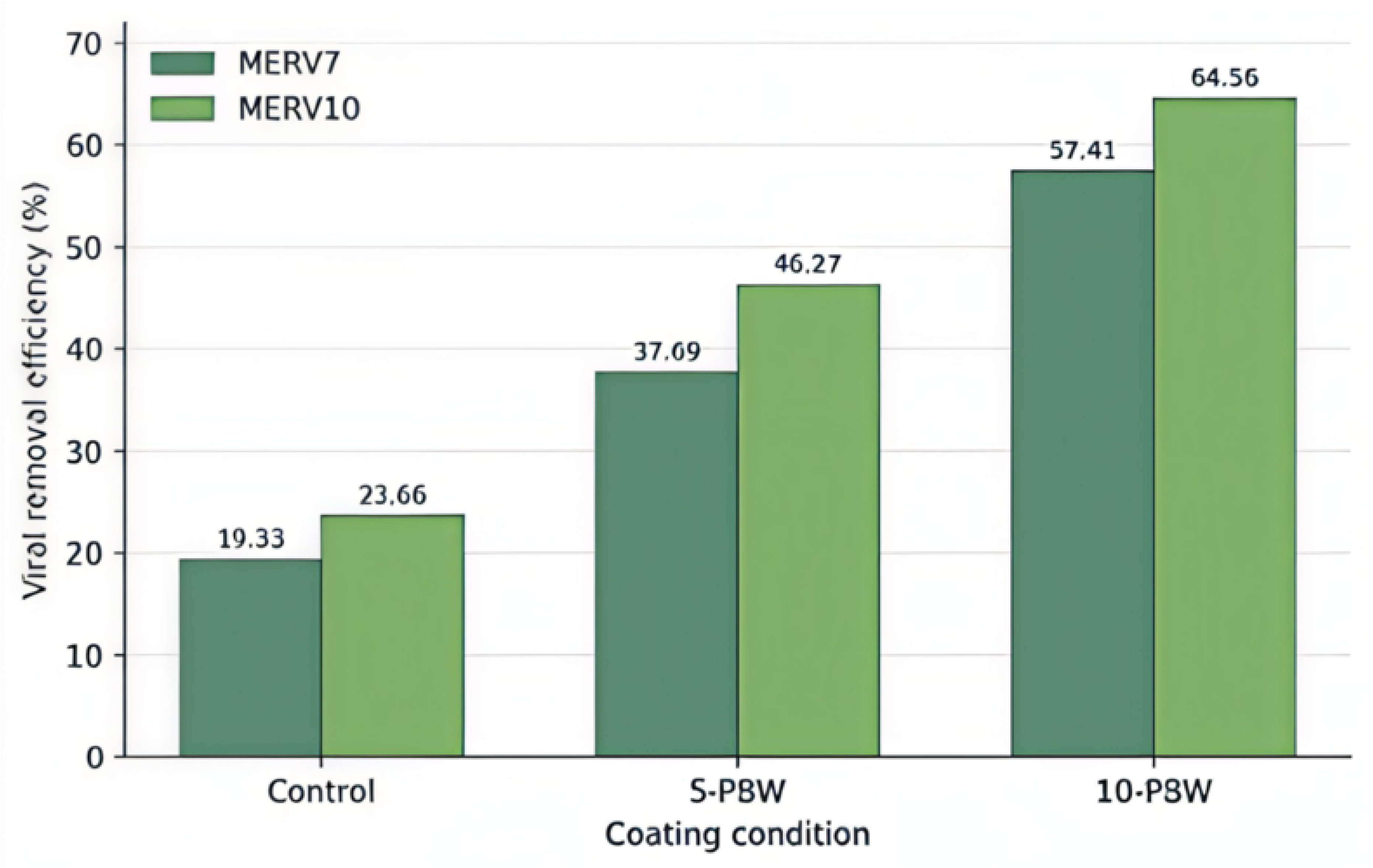
Antimicrobial coating technology demonstrates superior performance across multiple substrates, application methods, and MERV ratings. **a,** Viral filtration efficiency comparison across commercial filter substrates under high-flow ventilation conditions. 5W100 (MERV 8, left panel) and SP-100 (MERV 10, right panel) media were tested as untreated controls and with ablative polymer coating applied by spray and dip methods. SP-100 dipped media achieved 86.7% ± 1.31% VFE, exceeding MERV 15 performance thresholds (≥85% VFE) while maintaining MERV 10 pressure drop characteristics. Dipped application consistently showed superior performance across substrates. Data represent mean ± standard deviation (n=3), with all treated conditions showing statistically significant improvements (p < 0.001). **b,** Concentration-dependent viral filtration efficiency across MERV 7 and MERV 10 filter media. Both substrate types demonstrated progressive improvements with increasing ablative polymer concentration, validating scalability of the coating technology across standard commercial filtration media. Results demonstrate robust performance enhancement independent of baseline substrate MERV rating.

**Table 2:** Single-pass viral filtration efficiency results. Viral filtration efficiency testing conducted by LMS Technologies using MS2 bacteriophage challenge at 819 CFM under ASHRAE 52.2-compliant conditions. Data represent mean ± SD (n=3 per group) with statistical analysis by two-sample t-tests. SP-100 coated media achieved MERV 15+ performance (86.70% VFE) on MERV 10 substrates with minimal energy penalty (6.87% pressure increase). Extremely large effect sizes (Cohen’s d > 19) demonstrate robust, reproducible improvements. P-values < 0.001 indicate definitive statistical significance. Results represent first successful conversion of mechanical filtration substrates to high-efficiency biological performance through ablative coating technology.

| Medium | Treatment | VFE (%) | 95% CI | P-value* | Cohen's d | Pressure Drop | % Increase |
| --- | --- | --- | --- | --- | --- | --- | --- |
| SP-100 | Control | $30.80 \pm 0.79$ | 28.8-32.8 | - | - | 0.335 | - |
| SP-100 | Ablative coating | $86.70 \pm 1.31$ | 83.5-89.9 | **<0.001** | 51.68 | 0.358 | 6.87% |
| 5W100 | Control | 20.67 ±<br>1.36 | 17.3-24.0 | - | - | 0.110 | - |
| 5W100 | Ablative<br>coating | 52.87 ±<br>1.86 | 48.2-57.5 | <0.001 | 19.77 | 0.117 | 6.36% |

**Table 3:** Pressure drop analysis across filter substrates at 819 CFM. Independent pressure drop characterization across multiple commercial substrates demonstrates minimal aerodynamic penalties following ablative polymer coating. SP-100 substrate (used in viral testing) shows 5.6% pressure increase, validating the energy efficiency claims from biological performance testing.

| **Substrate** | **Control**<br>(in w.g.) | **Coated**<br>(in w.g.) | **Difference** | **%**<br>**Change** |
| --- | --- | --- | --- | --- |
| SP-100* | 0.310 | 0.327 | +0.017 | +5.6% |
| XP-100 | 0.275 | 0.316 | +0.041 | +14.9% |
| 1" PE | 0.116 | 0.128 | +0.012 | +10.3% |
| 2" PE | 0.180 | 0.172 | -0.008 | -4.4% |

**Table 4:** Summary of environmental emissions testing for control and treated filters at 400 cfm. Measured outlet and inlet concentrations of benzalkonium chloride, vinyl acetate, benzene, and benzyl chloride are shown for untreated and treated filter runs. All target analytes were at or below laboratory reporting limits. Among the broader TO-15 analyte library, only acetone, n-hexane, and cyclohexane were detected above reporting limits, and these occurred in both control and treated samples, consistent with room-air background.

| Compound | Units | Control |  | Filter |  | Flow Rate(CFM) |
| --- | --- | --- | --- | --- | --- | --- |
|  |  | Inlet | Outlet | Inlet | Outlet |  |
| Benzalkonium Chloride | µg/m<sup>3</sup> | <0.91 | <0.85 | <0.91 | <0.85 | 400 |
| Vinyl Acetate | ppbv | <0.50 | <0.50 | <0.50 | <0.50 | 400 |
| Benzene | ppbv | <0.85 | <0.85 | <0.85 | <0.85 | 400 |
| Benzyl Chloride | ppbv | <0.80 | <0.80 | <0.80 | <0.80 | 400 |

Simultaneous pressure drop analysis confirmed minimal energy penalties across multiple substrates (Table 3). SP-100 substrate showed only 5.6% pressure increase (0.310 to 0.327 in w.g.) at 819 CFM, validating previous observations that filter pressure drop significantly impacts HVAC system performance^28,29^. Coated media demonstrated substrate-dependent pressure changes ranging from beneficial reduction (−4.4% for 2" PE) to modest increases (14.9% for XP-100), confirming that biological performance improvements can be achieved without proportional energy penalties.

Concentration-dependent validation across MERV 7 and MERV 10 substrates demonstrated robust scalability of the coating technology (Figure 6b). Both substrate showed progressive VFE improvements with increasing coating concentration, confirming performance enhancement independent of baseline MERV rating. Energy-dispersive X-ray spectroscopy confirmed coating localization along fiber surfaces without disrupting porous filter architecture.

These results demonstrate successful conversion of MERV 10 mechanical substrates to MERV 15+ biological performance while maintaining MERV 10 pressure drop characteristics, representing the first demonstration of achieving high-efficiency biological performance through ablative coating without substantial energy penalties typically associated with passive filter upgrades.

Measurements conducted under standard conditions (70°F, 40% RH) using ASHRAE procedures. Negative values indicate beneficial pressure reductions. Results support implementation feasibility without substantial energy penalties typically associated with passive filter upgrades. The SP-100 results independently validate the 6.87% pressure increase observed in viral filtration testing, confirming minimal energy penalties for MERV 13+ biological performance. Notably, some substrates (2" PE) showed pressure drop reductions, indicating substrate-specific optimization potential.

### Environmental emissions testing found target analytes at or below reporting limits

Environmental testing showed that bacterial aerosol counts (BAC) and selected volatile organic compounds (VOCs) targets remained at or below laboratory reporting limits at both inlet and outlet locations for control and treated runs at 400 CFM. Outlet BAC was reported at <0.85 µg m⁻³ and inlet BAC at <0.91 µg m⁻³. Vinyl acetate was reported at <0.85 ppbv, benzene at <0.50 ppbv, and benzyl chloride at <0.80 ppbv at both inlet and outlet. Of the broader TO-15 analyte library, only acetone, n-hexane, and cyclohexane were detected above reporting limits, and these were present in both control and treated samples, consistent with room-air background contamination rather than emissions attributable to the treated filter

## Discussion

### Ablative Efficacy and Mechanistic Evidence

These results support the concept that ablative polymer-coated filtration media can convert passive fibrous substrates into active air-biofiltration materials that reduce downstream viral recovery while preserving practical airflow characteristics. The strongest evidence for this claim comes from the combination of concentration-dependent aerosol challenge performance, higher-flow single-pass filtration gains with small pressure-drop changes, and supportive TEM evidence of progressive structural disruption. The CFD results provide valuable mechanistic insights, though they should be framed as supportive rather than definitive. The modeled shift toward longer traversal behavior in the coated medium is consistent with increased opportunities for particle–surface interaction, but the reconstructed-geometry caveat means CFD cannot stand alone as proof of mechanism. However, when combined with the imaging and infectivity datasets, these results connect the modeled transport changes to observable viral structural damage and reduced downstream recovery of infectious virus. This integrated approach using computational fluid dynamics, microscopy, and biological assays follows established methodologies for studying particle transport and filtration in complex media[24,26].

### Mechanistic Considerations for BAC-Mediated Viral Inactivation

At the 1% BAC concentration used here, well above its critical micelle concentration (∼0.02%), BAC likely acts through micelle-mediated mechanisms rather than monomer adsorption^3**8**,3**9**^. Below the CMC, antiviral activity arises from stochastic BAC monomer adsorption to the negatively charged MS2 capsid³⁹. Above the CMC, the nonlinear inactivation increase suggests cooperative micelle-mediated mechanisms. The copolymer may enhance BAC activity by localizing micelles at the virus surface through combined hydrophobic and electrostatic interactions, leading to synergistic virucidal activity. Unlike conventional ablative coatings that suffer from leaching depletion or mechanical delamination, this system demonstrates non-leaching, surface-bound ablative activity. The polymer matrix likely provides sustained effectiveness through controlled surface renewal during aerosol exposure, maintaining an underlying reservoir of material bound to filter fibers rather than experiencing rapid depletion common in conventional ablative surfaces.

### Practical Implementation Advantages

The minimal pressure drop penalties (5.6-14.9% increases, with some beneficial reductions) while achieving substantial viral removal improvements distinguish this approach from conventional passive filter upgrades that pair biological gains with significant fan-energy burdens[33–36]. This pattern supports a different pathway to improved air-biofiltration performance than simply moving to denser passive media, addressing the well-documented relationship between filter pressure drop and HVAC system energy consumption[23]. This pattern supports a different pathway to improved air-biofiltration performance than simply moving to denser passive media. The combination of MERV 15+ biological performance on MERV 10 substrates, coupled with anticipated 50% pressure drop reduction in pleated configurations, positions this technology for retrofitting existing HVAC systems without substantial infrastructure modifications. This addresses documented challenges where high-efficiency filtration often exceeds existing HVAC capacity[35,36] while providing the indoor air quality benefits demonstrated in controlled environments[37]. The sustained antimicrobial activity and minimal energy penalties enable maintained effectiveness over extended operational periods without frequent replacement or increased energy costs.

### Environmental Safety and Emissions Profile

The emissions data strengthen the implementation case by showing that selected BAC-related and TO-15 analytes remained at or below reporting limits under tested conditions. These safety findings support the practical implementation of ablative filtration technology, complementing previous research on air cleaning device effectiveness[21] and addressing indoor air quality considerations crucial for occupied building applications[37].

### Study Limitations and Future Directions

This study focused on MS2 bacteriophage under controlled laboratory conditions, following established protocols for viral aerosol studies[15,20]. Future work should evaluate performance against envelope viruses (e.g., influenza), assess long-term durability under real-world HVAC conditions, and optimize coating formulations for different substrates and operational environments. Future mechanistic studies should employ cryo-electron microscopy and molecular dynamics simulations to elucidate specific molecular-level interactions between BAC micelles, the copolymer matrix, and viral capsids, providing direct evidence for the proposed synergistic inactivation mechanism. This work should build on the growing understanding of airborne pathogen transmission[22,38–40] and computational approaches for the optimization of filtration system [24,26,27].

## Author Contributions

Conceptualization: Ralph G. Dacey Jr., Chukwudi C. Ezeala, Robert Roth Data curation: Chukwudi C. Ezeala

Formal analysis: Emily Agnello, Alexander Ribbe, Sérgio Cavaleiro, Nelson Marques, Ralph G. Dacey Jr., Robert Roth

Investigation: Chukwudi C. Ezeala, Tom Kennedy, Robert Roth, Emily Agnello, Alexander Ribbe

Methodology: Chukwudi C. Ezeala, Tom Kennedy, Robert Roth

Project administration: Ralph G. Dacey Jr., Robert Roth

Resources: Ralph G. Dacey Jr., Tom Kennedy, Robert Roth

Software: Sérgio Cavaleiro, Nelson Marques

Supervision: Ralph G. Dacey Jr., Robert Roth

Validation: Chukwudi C. Ezeala, Emily Agnello, Alexander Ribbe

Visualization: Chukwudi C. Ezeala, Sérgio Cavaleiro, Nelson Marques

Writing – original draft: Ralph G. Dacey Jr

Writing – review & editing: All authors.

## Acknowledgments

We thank Nelson Laboratories for aerosol challenge testing, LMS Technologies for single-pass filtration analysis, FS Dynamics for computational fluid dynamics modeling, TRC Environmental Corporation for emissions testing, and the University of Massachusetts Lowell Institute for Applied Life Sciences for transmission electron microscopy analysis.

## Data Availability

All experimental data supporting the findings are provided in the Supporting Information files. The ablative coating formulation and application protocols are available to qualified researchers for academic purposes through material transfer agreements. Requests should be directed to the corresponding author or BioActive Technology LLC.

